# Cerebrospinal Fluid Myeloperoxidase Is Associated With Putamen Volume Beyond Neurofilament Light in Huntington’s Disease

**DOI:** 10.64898/2026.08.28.26361663

**Authors:** Jordan D. Clemsen, H. Jeremy Bockholt, William H. Adams, Bradley T. Baker, Jessica L. Bolton, Vince D. Calhoun, Jane S. Paulsen

**Author notes:** Corresponding Author: Jane S. Paulsen, Ph.D., University of Wisconsin–Madison, 7144 Medical Foundation Centennial Building, 1685 Highland Avenue, Madison, WI 53705, USA.

## Abstract

**Background:** The primary neuroanatomical site of Huntington’s disease (HD) pathology resides in the striatum and its atrophy identifies important disease progression from HD-ISS Stage 0 to Stage 1. Immune-associated proteins may capture variation in HD that is incompletely represented by markers of neuroaxonal injury.

**Objectives:** To determine whether cerebrospinal-fluid myeloperoxidase contributes information about striatal volume loss beyond genetic disease burden and neurofilament light.

**Methods:** Cross-sectional data from 88 persons with HD were analyzed. Cerebrospinal-fluid myeloperoxidase and neurofilament light were measured with a nucleic acid-linked immunosandwich assay. Normalized putamen volume was derived from structural magnetic resonance imaging. Linear regression adjusted for genetic disease burden and sex.

**Results:** Higher neurofilament light was associated with smaller normalized putamen volume (standardized β = −0.322, (P=.0066)). Higher myeloperoxidase was associated with larger normalized putamen volume after adjustment for genetic disease burden, sex, and neurofilament light (standardized β = 0.183, (P=.0386)). Adding myeloperoxidase increased explained variance in striatal loss.

**Conclusions:** Cerebrospinal fluid myeloperoxidase contributed modest incremental information about striatal volume in this cross-sectional sample. Independent longitudinal studies are needed to determine its biological source, temporal behavior, and potential biomarker value. Findings advance efforts to characterize multicomponent biological markers of HD.

## Introduction

Huntington’s disease (HD) is an autosomal dominant neurodegenerative disorder caused by an expanded CAG repeat in the *HTT* gene, resulting in mutant huntingtin protein (mHTT) with an expanded polyglutamine tract.^1^ Although the genetic cause is established, the downstream processes contributing to neurodegeneration remain incompletely understood.^2, 3^ While the general understanding is that mHTT influences HD progression, the specific mechanism(s) for neurodegeneration remains unknown.^4^ Despite having reliable measurements of the pathogenic protein,^5–9^ biomarker efforts are ongoing to elucidate the pathogenesis of the disease. Currently, persons with HD share a genetic burden associated with age at disease onset, most typically defined by a CAG-Age-Product (CAP) score,^10^ though findings are mixed on the association of CAP with disease progression rate. Recent work has emphasized several biomarkers beyond genetic burden alone,^11–15^ including Neurofilament Light Chain (NEFL) and Tumor Necrosis Factor Receptor Superfamily member 8 (TNFRSF8), providing the basis for a two-track model.^16^ In this work, cerebrospinal fluid (CSF) proteomics identified biofluid measures improving accuracy of disease prognosis defined by striatal volumes. Mediation analysis suggested an immune-associated axis independent of pathological cytoskeletal damage. However, TNFRSF8 is involved in multiple pathways and biological processes that may not influence disease progression. While the two-track model involving NEFL and TNFRSF8 demonstrates some underlying HD features by incorporating downstream innate immune activation of biomarker candidates, extending the model could provide additional insight into the early HD stages.^16, 17^ TNFRSF8 belongs to the TNF receptor superfamily expressed predominantly by activated lymphocytes and participates in receptor-mediated immune regulation. Myeloperoxidase (MPO) captures a biologically distinct feature of the immune response. This heme peroxidase is released primarily by activated neutrophils and other myeloid cells and uses hydrogen peroxide and halides to generate reactive oxidants, including hypochlorous acid. Its abundance may therefore provide a marker of innate myeloid activation and oxidative inflammatory activity. MPO has been implicated in vascular inflammation, ischemic brain injury, and neurodegenerative pathology, with increased MPO expression or immunoreactivity reported in affected brain regions in Parkinson’s and Alzheimer’s disease.^18–20^ Evidence in HD remains limited. We examined CSF MPO to determine whether the Track-2 signal generalizes beyond TNFRSF8/CD30 and encompasses a broader immune-associated process related to striatal integrity. We therefore tested whether MPO explained additional variance in normalized putamen volume (nPV) after accounting for genetic disease burden, sex, and neuroaxonal injury reflected by NEFL.

## Methods

### Participants and Measures

Participants from the Predict-HD and Prevent-HD observational studies were included if they had available CSF proteomics data, structural magnetic resonance imaging (MRI)-derived volumetric measures, and disease burden metrics based on CAG repeat length and age at the study visit. In these studies, participants completed multiple clinical examinations from baseline up to 12 years. During visits, CSF from willing participants was collected by lumbar puncture after overnight fasting. Immediately following extraction, CSF was subject to quality control, then centrifuged to be stored at the NINDS BioSEND Repository for centralized data sharing. Primary models used complete-case data for nPV, CAP score, sex, NEFL, and MPO.

Research protocols were approved by the University of Iowa and University of Wisconsin (UW) Institutional Review Boards (IRB# 200901701, 201301724, 2020-1254) and at each site. All participants gave written informed consent. Both studies were registered on ClinicalTrials.gov (NCT00051324, NCT04818060). All phenotype, imaging, and biosample data are shared through the National Data Archive, the NIH database of Genotypes and Phenotypes (dbGaP under accession phs000222.v6.p2), BioSEND, and the authors’ labs. We confirm that all ethical and regulatory requirements have been met and that the research was conducted in accordance with the principles of the Declaration of Helsinki and applicable institutional policies. This research used de-identified participant data. Data access is controlled and requires submission of a data request to dbGaP, approval by the relevant Data Access Committee, and execution of a Data Use Agreement with UW.

Disease burden, coined by Penney and colleagues, was calculated based on neuropathology samples from known HD participants.^21^ The formula was updated in the Predict-HD study to better represent earlier stages of the disease course prior to death.^22, 23^ CAG repeat length is the number of trinucleotide repeats in the *HTT* gene. The CAP score developed from early Predict-HD clinical data is computed as CAP = (Age at Study Entry) ⋅ (CAG – L) / K where L is a constant CAG length and K is a scaling constant. The CAP score has been used fieldwide as a continuous or categorical measure to represent the cumulative disease burden for living persons carrying the gene expansion before clinical motor diagnosis.^10^

The HD Integrated Staging System (HD-ISS) is a staging model that assigns an ordinal value based on disease severity with Stage 0 representing Individuals with ≥ 40 CAG repeats in the *HTT* gene and no biological signs or clinical symptoms.^24^ Stage 1 represents detectable progression of HD with measurable indicators of underlying pathophysiology (defined using MRI measures). Stage 2 represents detectable clinical phenotype, expressed as quantified scores on the Symbol Digit Modality Test (SDMT) and the Total Motor Score (TMS).^25, 26^ Stage 3 represents a decline in function where individuals may experience significant impairment delineated with the Total Functional Capacity (TFC) scale.^25, 27^ Though the HD-ISS was developed to facilitate the design of clinical trials targeting populations before clinical motor diagnosis and to enable data standardization across ongoing and future studies, the potential biological markers validated for stage 1 need to be identified and elucidated in a standardized manner across datasets. Because HD-ISS Stage 1 is defined by detectable neurodegeneration prior to overt clinical manifestations, biomarkers reflecting complementary biological processes may improve biological characterization during this earliest disease stage.^24, 28^

### CSF Proteomics

CSF proteomic profiling was performed at the UW Alzheimer’s Disease Research Center Biomarker Laboratory using the NULISAseq™ CNS Disease Panel 120 and NULISAseq™ Inflammation Panel 250. The two panels measure proteins related to neurodegeneration, synaptic function, and inflammatory and immune signaling. NEFL was measured on the CNS Disease Panel 120; MPO was measured on the Inflammation Panel 250. Assays were performed on the Alamar ARGO HT System according to the manufacturer’s protocol by laboratory personnel blinded to participant characteristics and imaging results.

Nucleic acid-linked immunosandwich assay (NULISA) uses paired target-specific capture and detection antibodies conjugated to unique oligonucleotide tags. Following incubation with CSF, the resulting immunocomplexes undergo sequential magnetic-bead capture, washing, release, and recapture to remove unbound antibodies and reduce background signal. Binding of both antibodies to the target protein brings their oligonucleotide tags into proximity for ligation. The resulting target- and sample-specific DNA reporters are amplified and quantified by next-generation sequencing using the Illumina NextSeq 1000 System.

Counts for each analyte were first normalized to the internal control within each sample well. The internal-control-normalized counts were then divided by the analyte-specific median of the interplate control replicates on the same plate. The resulting values were rescaled and log2 transformed to generate NULISA Protein Quantification (NPQ) values.^22^ NPQ represents relative protein abundance. Values can be compared across participants for the same analyte; NPQ values from different analytes are not directly comparable.

Reported NEFL and MPO values were selected a priori as Track 1 and Track 2 candidates, respectively, and were examined for assay quality flags, detection frequency, and extreme observations before statistical analysis. For the current study, NEFL was treated as the primary neuroaxonal injury marker; MPO was selected to test an orthogonal immune/oxidative mechanism relative to TNFRSF8/CD30 findings previously published.^16^

### Neuroimaging Outcomes

Predict-HD neuroimaging methods have been described in detail elsewhere.^29^ In brief, T1-weighted structural MRI scans were collected on GE scanners from multiple sites internationally. For Prevent-HD, participants completed 60-minute MRI sessions at baseline and 18-month follow-up on a 3T GE scanner at a single site to collect estimates of brain and CSF volume, structure, diffusion tensor estimates, functional brain connectivity data, and arterial spin labeling data. For both studies, caudate and putamen were normalized by intra-cranial volume. Given that HD pathogenesis involves variations in neurodevelopment as well as neurodegeneration and that the wild HTT influence on the putamen is noticeable before the caudate in previous MRI studies of HD before clinical motor diagnosis, this analysis used the putamen as the primary region of interest.^30^ Data were organized according to the Brain Imaging Data Structure standard.^31^ Raw DICOM files collected from the Predict-HD study were redistributed using the FastSurfer atlas.^32^ Manual quality control was applied to subject DICOM data to account for ghost artifacts and other motion, misalignment, and acquisition issues. In the regional MRI screening, nPV demonstrated the strongest association with CAP score (*r* = −0.64; *p* < .001), validating it as the primary neuroimaging outcome.

### Statistical Methods

Participants’ quantitative characteristics are summarized using means with standard deviations (SD) as well as medians with interquartile and full ranges. Categorical characteristics are summarized using frequencies and percentages. The primary outcome was nPV. Primary predictors of interest were NEFL and MPO. Age, sex, and CAP score were evaluated as demographic and disease-burden covariates. Three MPO measurements (3.4%) were below the assay limit of detection and assigned a value of zero by the NPQ analysis pipeline.

Associations with nPV were evaluated using ordinary least-squares linear regression. Univariable models were first fit separately for sex, NEFL, MPO, age, and CAP score. Three multivariable models were subsequently evaluated. Model 1 included sex, NEFL, and MPO. Model 2 additionally included age. Model 3 included sex, NEFL, MPO, and CAP score. Regression results are presented as unstandardized regression coefficients (β) with 95% confidence intervals (CIs), standardized regression coefficients, and *p*-values. CAP coefficients are presented per 10-unit increase. Model fit was summarized using R², adjusted R², Akaike’s information criterion (AIC), and the Bayesian information criterion (BIC), with lower AIC and BIC values indicating better relative fit.

To quantify the incremental contribution of MPO in the CAP-adjusted model, Model 3 was compared with a reduced model containing CAP score, sex, and NEFL. The change in R² associated with the addition of MPO was calculated, and the incremental contribution of MPO was evaluated using a partial *F*-test. For all models, the assumptions of linearity, normality, and homoscedasticity were assessed using residual plots, influential observations were assessed using Cook’s distance, and multicollinearity was evaluated using variance inflation factors.

Exploratory indirect-effect analyses were conducted to evaluate two alternative statistical pathways linking CAP score, MPO, and nPV. Model 1 specified CAP → MPO → nPV; Model 2 specified CAP → nPV → MPO. Indirect effects were calculated as the product of the *a* and *b* path coefficients. Confidence limits (95% CI) for the indirect effects were estimated using 5,000 nonparametric bootstrap resamples and the percentile method. The Sobel *z*-statistic and corresponding *p*-value were additionally calculated using the first-order Sobel approximation. These analyses were unadjusted. Because all measurements were cross-sectional, the specified pathways were considered alternative statistical models and not interpreted as establishing temporal or causal direction.

Associations of NEFL and MPO with nPV were additionally visualized using scatterplots with ordinary least-squares regression lines and 95% CIs for the mean regression line. All statistical tests were two-sided, with *p* < .05 considered statistically significant. Statistical analyses were performed using Python and SAS version 9.4 (Cary, NC).

## Results

### Participant Characteristics

The sample included 88 participants with a mean age of 39.0 (SD = 11.8) years. Fifty-eight (65.9%) participants were female and 30 (34.1%) were male. Most participants were White (n = 87 or 98.9%) and non-Hispanic (n = 84 or 95.5%). Participants had a mean of 42.7 (SD = 2.9) CAG repeats and a mean CAP score of 336.5 (SD = 92.3). The mean TMS was 6.5 (SD = 9.7), mean TFC was 12.3 (SD = 1.6), and mean SDMT score was 55.2 (SD = 14.2). The mean NEFL and MPO values were 16.5 (SD = 1.1) and 4.2 (SD = 1.2), respectively. The mean nPV was 0.7 (SD = 0.1) (**Table 1**).

**Table 1:** Participant characteristics.

*Participant characteristics*
|  | Total<br>(N=88) |
| --- | --- |
| **Age** |  |
| Mean (SD) | 39.0 (11.8) |
| Median (IQR) | 36.4 (29.8, 46.3) |
| Median (Range) | 36.4 (19.8, 68.1) |
| **Years of education** |  |
| Missing | 3 |
| Mean (SD) | 15.3 (2.2) |
| Median (IQR) | 16.0 (14.0, 16.0) |
| Median (Range) | 16.0 (11.0, 20.0) |
| **Gender, n (%)** |  |
| Female | 58 (65.9%) |
| Male | 30 (34.1%) |
| **Race, n (%)** |  |
| White | 87 (98.9%) |
| Other/Not Reported | 1 (1.1%) |
| **Ethnicity, n (%)** |  |
| Non-Hispanic | 84 (95.5%) |
| Hispanic | 3 (3.4%) |
| Other/Not Reported | 1 (1.1%) |
| **Cytosine-adenine-guanine (CAG) repeats** |  |
| Mean (SD) | 42.7 (2.9) |
| Median (IQR) | 42.0 (41.0, 44.0) |
| Median (Range) | 42.0 (37.0, 54.0) |
|  | Total<br>(N=88) |
| **CAG-by-age product (CAP) score** |  |
| Mean (SD) | 336.5 (92.3) |
| Median (IQR) | 340.0 (265.3, 380.2) |
| Median (Range) | 340.0 (160.0, 659.0) |
| **HD-ISS Stage, n (%)** |  |
| 0 | 42 (50.6%) |
| 1 | 19 (22.9%) |
| 2 | 15 (18.1%) |
| 3 | 7 (8.4%) |
| Missing | 5 |
| **Total Motor Score** |  |
| Missing | 3 |
| Mean (SD) | 6.5 (9.7) |
| Median (IQR) | 4.0 (0.0, 9.0) |
| Median (Range) | 4.0 (0.0, 54.0) |
| **Total Functional Capacity** |  |
| Missing | 3 |
| Mean (SD) | 12.3 (1.6) |
| Median (IQR) | 13.0 (13.0, 13.0) |
| Median (Range) | 13.0 (5.0, 13.0) |
| **Symbol Digit Modalities Test** |  |
| Missing | 4 |
| Mean (SD) | 55.2 (14.2) |
| Median (IQR) | 54.0 (46.5, 63.0) |
| Median (Range) | 54.0 (22.0, 110.0) |
| **Neurofilament light chain** |  |
| Mean (SD) | 16.5 (1.1) |
| Median (IQR) | 16.6 (15.8, 17.3) |
| Median (Range) | 16.6 (13.6, 18.5) |
| **Myeloperoxidase** |  |
| Mean (SD) | 4.2 (1.2) |
| Median (IQR) | 4.4 (3.8, 4.9) |
| Median (Range) | 4.4 (0.0, 5.9) |
| **Normalized putamen volume** |  |
| Mean (SD) | 0.7 (0.1) |
| Median (IQR) | 0.7 (0.6, 0.8) |
|  | Total<br>(N=88) |
| Median (Range) | 0.7 (0.4, 1.4) |
| *Note: Three myeloperoxidase (MPO) measurements (3.4%) were below the assay limit of detection and were assigned a value of 0 by the Nucleic Acid Linked Immuno-Sandwich Assay (NULISA) Protein Quantification (NPQ) analysis pipeline. |  |

### Associations With nPV

In univariable analyses, higher NEFL was associated with lower nPV (β = −0.07; 95% CI, −0.09 to −0.04; standardized β = −0.53; *p* < .001) whereas higher MPO was associated with higher nPV (β = 0.03; 95% CI, 0.01 to 0.05; standardized β = 0.27; *p* = .011). Higher CAP score was also associated with lower nPV (β per 10-unit increase = −0.008; 95% CI, −0.010 to −0.005; standardized β = −0.52; *p* < .001). Neither age nor sex was significantly associated with nPV in univariable analyses (**Table 2**).

**Table 2:**
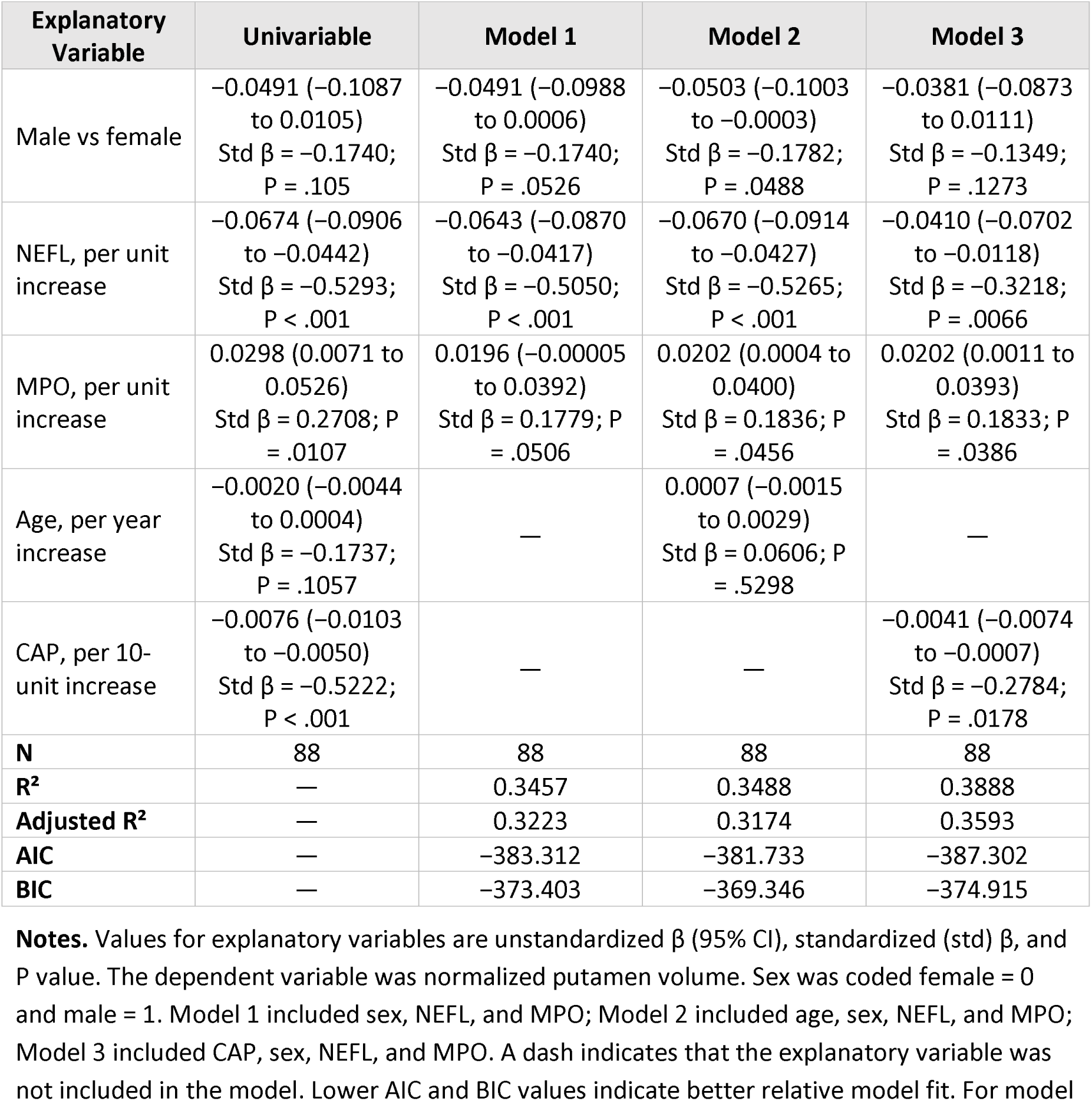
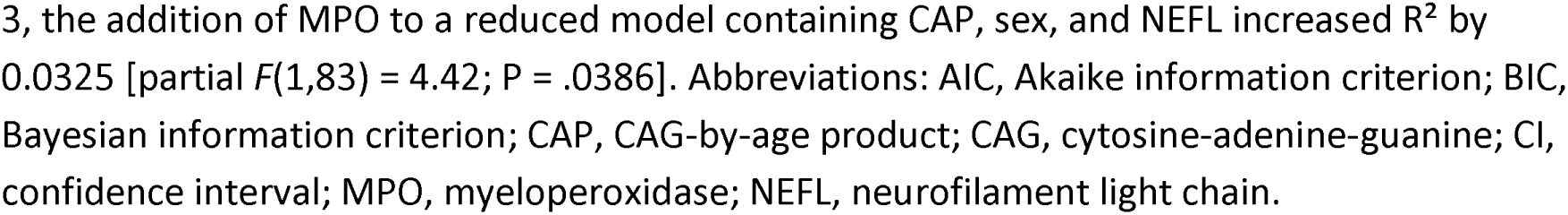
Multivariable regression models of normalized putamen volume incorporating neuroaxonal, immune, and disease-burden measures.

| Explanatory Variable | Univariable | Model 1 | Model 2 | Model 3 |
| --- | --- | --- | --- | --- |
| Male vs female | −0.0491 (−0.1087 to 0.0105)<br>Std $\beta$ = −0.1740;<br>P = .105 | −0.0491 (−0.0988 to 0.0006)<br>Std $\beta$ = −0.1740;<br>P = .0526 | −0.0503 (−0.1003 to −0.0003)<br>Std $\beta$ = −0.1782;<br>P = .0488 | −0.0381 (−0.0873 to 0.0111)<br>Std $\beta$ = −0.1349;<br>P = .1273 |
| NEFL, per unit increase | −0.0674 (−0.0906 to −0.0442)<br>Std $\beta$ = −0.5293;<br>P < .001 | −0.0643 (−0.0870 to −0.0417)<br>Std $\beta$ = −0.5050;<br>P < .001 | −0.0670 (−0.0914 to −0.0427)<br>Std $\beta$ = −0.5265;<br>P < .001 | −0.0410 (−0.0702 to −0.0118)<br>Std $\beta$ = −0.3218;<br>P = .0066 |
| MPO, per unit increase | 0.0298 (0.0071 to 0.0526)<br>Std $\beta$ = 0.2708; P = .0107 | 0.0196 (−0.00005 to 0.0392)<br>Std $\beta$ = 0.1779; P = .0506 | 0.0202 (0.0004 to 0.0400)<br>Std $\beta$ = 0.1836; P = .0456 | 0.0202 (0.0011 to 0.0393)<br>Std $\beta$ = 0.1833; P = .0386 |
| Age, per year increase | −0.0020 (−0.0044 to 0.0004)<br>Std $\beta$ = −0.1737;<br>P = .1057 | — | 0.0007 (−0.0015 to 0.0029)<br>Std $\beta$ = 0.0606; P = .5298 | — |
| CAP, per 10-unit increase | −0.0076 (−0.0103 to −0.0050)<br>Std $\beta$ = −0.5222;<br>P < .001 | — | — | −0.0041 (−0.0074 to −0.0007)<br>Std $\beta$ = −0.2784;<br>P = .0178 |
| N | 88 | 88 | 88 | 88 |
| R<sup>2</sup> | — | 0.3457 | 0.3488 | 0.3888 |
| Adjusted R<sup>2</sup> | — | 0.3223 | 0.3174 | 0.3593 |
| AIC | — | −383.312 | −381.733 | −387.302 |
| BIC | — | −373.403 | −369.346 | −374.915 |

**Notes.** Values for explanatory variables are unstandardized $\beta$ (95% CI), standardized (std) $\beta$ , and P value. The dependent variable was normalized putamen volume. Sex was coded female = 0 and male = 1. Model 1 included sex, NEFL, and MPO; Model 2 included age, sex, NEFL, and MPO; Model 3 included CAP, sex, NEFL, and MPO. A dash indicates that the explanatory variable was not included in the model. Lower AIC and BIC values indicate better relative model fit. For model
3, the addition of MPO to a reduced model containing CAP, sex, and NEFL increased $R^2$ by 0.0325 [partial $F(1,83) = 4.42$ ; $P = .0386$ ]. Abbreviations: AIC, Akaike information criterion; BIC, Bayesian information criterion; CAP, CAG-by-age product; CAG, cytosine-adenine-guanine; CI, confidence interval; MPO, myeloperoxidase; NEFL, neurofilament light chain.

In Model 1, which included sex, NEFL, and MPO, NEFL remained inversely associated with nPV (β = −0.06; 95% CI, −0.09 to −0.04; standardized β = −0.51; *p* < .001). The MPO association was attenuated after adjustment (β = 0.02; 95% CI, −0.0001 to 0.04; standardized β = 0.18; *p* = .051). Model 1 explained 34.6% of the variance in nPV (R² = .346; adjusted R² = .322).

After additionally adjusting for age in Model 2, NEFL remained inversely associated with nPV (β = −0.07; 95% CI, −0.09 to −0.04; standardized β = −0.53; *p* < .001), and MPO was positively associated with nPV (β = 0.02; 95% CI, 0.0004 to 0.04; standardized β = 0.18; *p* = .046). Age remained unassociated with nPV after adjusting for sex, NEFL, and MPO (β = 0.001; 95% CI, −0.002 to 0.003; *p* = .53). Model 2 had R² = .349 and adjusted R² = .317.

In Model 3, which included CAP score, sex, NEFL, and MPO, both biomarkers remained independently associated with nPV. Higher NEFL was associated with lower nPV (β = −0.04; 95% CI, −0.07 to −0.01; standardized β = −0.32; *p* = .01) whereas higher MPO was associated with higher nPV (β = 0.02; 95% CI, 0.001 to 0.04; standardized β = 0.18; *p* = .04). Higher CAP score was independently associated with lower nPV (β per 10-unit increase = −0.004; 95% CI, −0.01 to −0.001; standardized β = −0.28; *p* = .02). Sex was not independently associated with nPV in Model 3 (*p* = .13).

Model 3 explained 38.9% of the variance in nPV (R² = .389; adjusted R² = .359) and had the lowest AIC and BIC of the three multivariable models (AIC = −387.30; BIC = −374.92). Addition of MPO to the reduced model containing CAP score, sex, and NEFL increased R² by .033. This incremental contribution was statistically significant [partial F(1, 83) = 4.42; *p* = .04].

Consistent with the regression analyses, graphical examination demonstrated an inverse association between NEFL and nPV and a positive association between MPO and nPV (Figure 1).

**Figure 1.**
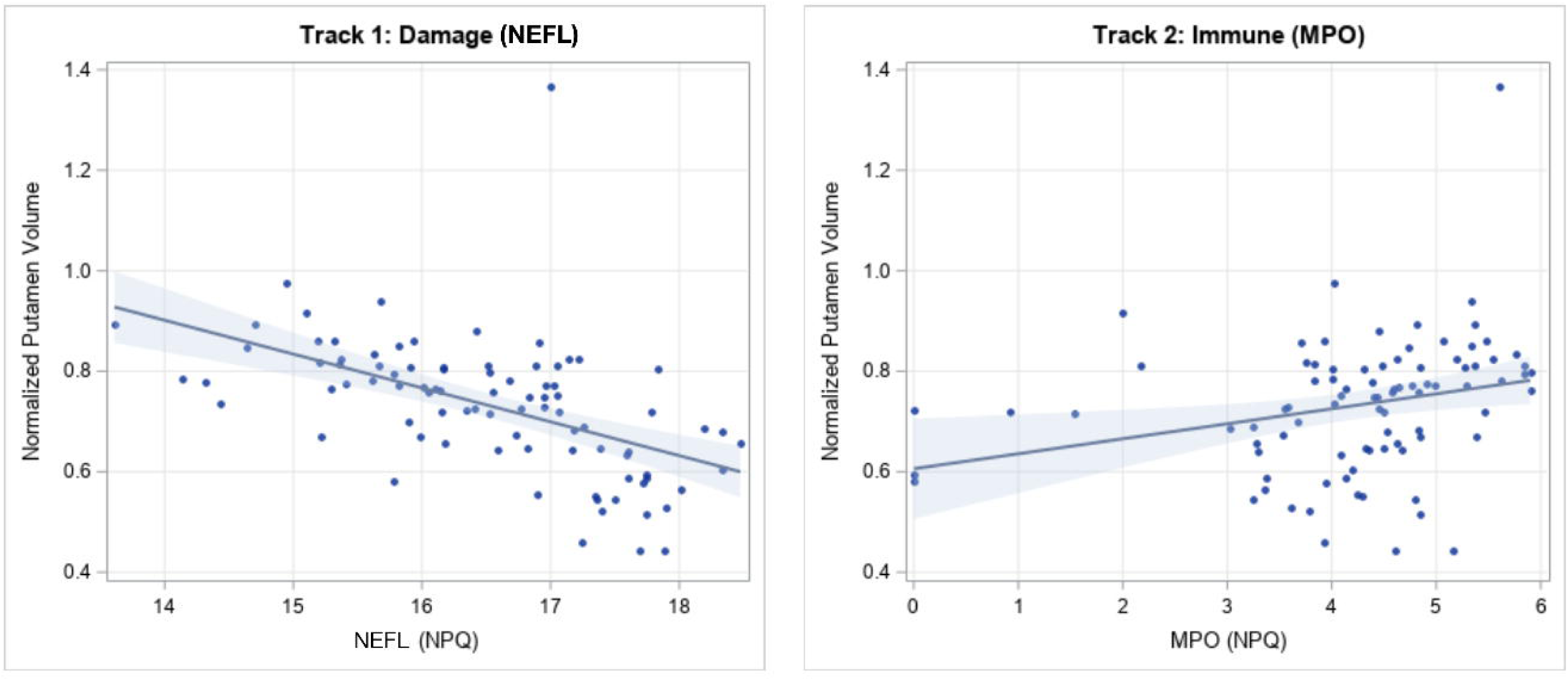
Associations of neurofilament light chain and myeloperoxidase with normalized putamen volume. Scatterplots show the relationships between normalized putamen volume and cerebrospinal fluid neurofilament light chain (NEFL; Track 1) and myeloperoxidase (MPO; Track 2). Solid lines represent ordinary least-squares regression fits, and shaded bands represent 95% confidence intervals for the mean regression line. NEFL was inversely associated with normalized putamen volume (Pearson r = −0.53, 95% CI: −0.66 to −0.36; p < .001) whereas MPO showed a positive association with normalized putamen volume (Pearson r = 0.27, 95% CI: 0.06 to 0.45; p = .01). NPQ indicates NULISA Protein Quantification.

### Sensitivity Analysis of Below-Limit MPO Measurements

In a sensitivity analysis excluding three participants with MPO measurements below the assay limit of detection, the magnitude and direction of the MPO association in Model 3 were like the primary analysis (β = 0.02; 95% CI, −0.002 to 0.05) although the association was no longer statistically significant (*p* = .07).

### Exploratory Indirect-Effect Analyses

In the exploratory analysis specifying CAP → MPO → nPV (Model 1), the estimated indirect effect was −0.00003 (95% bootstrap CI, −0.0001 to 0.00002). The confidence interval included zero. The corresponding Sobel test was also nonsignificant (*z* = −0.84; *p* = .40).

In the alternative model specifying CAP → nPV → MPO (Model 2), the estimated indirect effect was −0.002 (95% bootstrap CI, −0.005 to −0.001), with the bootstrap confidence interval excluding zero. The corresponding Sobel test was also statistically significant (*z* = −2.28; *p* = .02) (Table 3). These findings indicate that the observed cross-sectional associations were consistent with an indirect association under the second specified statistical ordering but not the first. Because the data are cross-sectional, the analyses do not, however, establish the temporal or causal ordering of nPV and MPO.

**Table 3:** Exploratory indirect-effect analyses of alternative pathways linking CAP, MPO, and normalized putamen volume.

| Model | Path A | Path B | Indirect Effect | 95% Bootstrapped CI |  | Sobel z | Sobel p |
| --- | --- | --- | --- | --- | --- | --- | --- |
|  |  |  |  | Lower | Upper |  |  |
| 1 | -0.001260 | 0.024560 | -0.000031 | -0.000111 | 0.000019 | -0.84 | 0.40 |
| 2 | -0.000762 | 2.757130 | -0.002100 | -0.004793 | -0.000861 | -2.28 | 0.02 |

Note: The specified pathway for Model 1 was CAP → MPO → normalized putamen volume; the specified pathway for Model 2 was CAP → normalized putamen volume → MPO. Indirect effects were calculated as the product of the $a$ and $b$ path coefficients. The 95% CIs were estimated using 5,000 nonparametric bootstrap resamples and the percentile method. The Sobel $z$ statistic and $p$ -value were calculated using the first-order Sobel approximation.

## Discussion

In this cross-sectional study of persons with HD, CSF NEFL and MPO demonstrated distinct associations with nPV after accounting for disease burden and other covariates. Higher NEFL was associated with lower nPV whereas higher MPO was associated with higher nPV. Both associations persisted after adjusting for CAP score and sex: the final model explained approximately 39% of the variance in nPV. Moreover, the addition of MPO to a model containing CAP score, sex, and NEFL increased the explained variance in nPV by 3.3%. Together, these findings suggest that NEFL and MPO provide distinct and potentially complementary information about structural variation in HD and may reflect different underlying biological processes rather than interchangeable markers of disease severity.

The inverse association between NEFL and nPV is consistent with the established role of NEFL as a marker of neuroaxonal injury.^16, 33^ Higher NEFL was strongly associated with lower nPV in univariable analysis and remained independently associated with lower nPV after adjusting for CAP score, sex, and MPO, indicating that this relationship was not fully accounted for by genetic disease burden.

In contrast, higher MPO was associated with greater nPV and remained positively associated after adjusting for CAP score, sex, and NEFL. This positive association is notable when considering MPO as a candidate marker of neuroinflammatory activity. Within these cross-sectional data, higher MPO did not correspond to greater putaminal atrophy. Rather, higher MPO concentrations were associated with greater nPV after accounting for measured disease burden and NEFL. This pattern raises the possibility that MPO reflects a biological process that may vary with disease stage or in relation to structural degeneration rather than functioning simply as a marker of cumulative neuronal injury.

MPO is an innate immune enzyme involved in oxidative inflammatory responses and is released during activation of neutrophils and other myeloid cells. Through reactions involving hydrogen peroxide, MPO generates reactive oxidants, including hypochlorous acid, that contribute to host defense but may also promote oxidative tissue injury under conditions of sustained or dysregulated inflammatory activity. Although MPO has been less extensively studied than NEFL in HD, its established relationship with innate immune activation and oxidative stress provides a biologically plausible basis for evaluation as a marker of processes distinct from neuroaxonal injury.^34, 35^ Importantly, the present findings do not establish whether MPO activity contributes directly to structural change or instead reflects an immune response occurring in association with neurodegeneration.

The present findings also extend previous work evaluating TNFRSF8/CD30 as an immune-associated biomarker within the Two-Track framework.^16^ Whereas TNFRSF8 reflects immune-regulatory signaling, MPO represents a distinct aspect of innate immune and oxidative activity. The observation that MPO remained associated with nPV after accounting for NEFL and CAP score suggests that the previously observed immune-associated signal may not be specific to a single biomarker or pathway. These findings raise the possibility that multiple immune-associated markers may provide complementary information to measures of neuroaxonal injury. Direct comparison or joint modeling of MPO and TNFRSF8, together with replication in independent cohorts, will be necessary to determine whether these markers reflect a common immune-associated process or distinct complementary biological pathways.

The exploratory indirect-effect analyses provide additional context for these findings. No indirect effect was detected under the statistical ordering CAP → MPO → nPV whereas the alternative ordering CAP → nPV → MPO yielded an indirect effect whose bootstrap confidence interval excluded zero. Thus, the observed cross-sectional relationships were consistent with an indirect association between CAP-related disease burden and MPO through variation in nPV. However, these analyses cannot establish that structural degeneration temporally precedes changes in MPO, nor can they exclude reciprocal relationships, common upstream mechanisms, or other unmeasured processes. Serial structural MRI and MPO measurements will be necessary to determine the temporal relationship between neurodegeneration and inflammatory activity.

These findings should also take into account assay measurement at the lower end of its range. Three participants had MPO measurements below the assay limit of detection and were assigned a value of zero by the NULISA NPQ analysis pipeline. Exclusion of these observations yielded an MPO coefficient similar in magnitude and direction to that of the primary analysis although the confidence interval widened and the association was no longer statistically significant. Thus, the positive association observed in the primary analysis does not appear to be explained solely by the below-limit measurements. However, the reduced precision of the sensitivity estimate underscores the need for confirmation in a larger sample.

Several limitations should be considered. First, the cross-sectional design precludes inference regarding temporal or causal relationships among disease burden, structural brain changes, and biomarker concentrations. This limitation is particularly important when interpreting the exploratory indirect-effect analyses. Second, the modest sample size limits precision for smaller associations and more complex multivariable models and may contribute to instability of estimates in sensitivity analyses. Third, residual confounding by unmeasured factors that may influence MPO concentrations, including medications, comorbid conditions, or systemic inflammatory activity, cannot be excluded. Finally, assigning values of zero to MPO measurements below the assay limit of detection introduces potential measurement uncertainty even though exclusion of these observations in sensitivity analysis yielded a similar effect estimate.

Despite these limitations, the findings support the potential value of evaluating markers of neuroaxonal injury and immune-associated activity together when characterizing HD-related structural variation. Whereas NEFL demonstrated the expected inverse association with nPV, MPO showed a distinct positive association after accounting for CAP score, sex, and NEFL. Rather than establishing a directional pathway between inflammation and neurodegeneration, these findings identify a potentially informative relationship between MPO and structural disease features that warrants further investigation. If replicated in independent cohorts, biomarkers reflecting complementary neuroaxonal and immune-associated processes may improve biological characterization beyond that provided by genetic disease burden alone. Longitudinal studies incorporating repeated biomarker measurements and serial neuroimaging will be necessary to determine whether MPO changes precede, accompany, or follow structural degeneration, whether their relationship with structural change varies across disease stages, and whether they ultimately have utility for biological stratification in HD.

## Data Availability

Data access is controlled and requires submission of a data request to dbGaP, approval by the relevant Data Access Committee, and execution of a Data Use Agreement with University of Wisconsin.

## Acknowledgment

The Predict-HD study was supported by NINDS/NIH and CHDI Foundation funding: “Neurobiological Predictors of Huntington’s Disease” (R01NS040068; R01NS082089); CHDI Grants A3917 and A6266. The PREVENT-HD study was supported by NINDS/NIH funding: “Statistical disease modeling and clinimetrics to prepare for preventative trials in Huntington’s disease” (U01NS103475); “Preparing for preventative clinical trials in Huntington’s disease” (U01NS105509). We appreciate the 1500 Predict-HD participants. We acknowledge the excellent collaborators in the Predict-HD and Prevent-HD studies. A full list is recorded here: https://neurology.wisc.edu/wp-content/uploads/2023/05/PREDICT-HD_2020_Acknowledgments.pdf; https://neurology.wisc.edu/wp-content/uploads/2026/08/PREVENT-HD_Acknowledgments.pdf. We appreciate administrative assistance from Deven K. Burks.

## Authors’ Roles

JDC: design, execution, analysis, writing; HJB: design, execution, analysis, editing final version of the manuscript; WHA: analysis, editing final version of the manuscript; BTB: analysis, editing final version of the manuscript; JLB: editing final version of the manuscript; VDC: editing final version of the manuscript; JSP: design, execution, writing.

## References

1. Aronin N, Chase K, Young C, et al. CAG expansion affects the expression of mutant Huntingtin in the Huntington’s disease brain. Neuron 1995;15(5):1193–1201.

2. Ghosh R, Tabrizi SJ. Clinical Features of Huntington’s Disease. In: Nóbrega C, Pereira de Almeida L, eds. Polyglutamine Disorders. Cham: Springer International Publishing, 2018:1–28.

3. Roos RA. Huntington’s disease: a clinical review. Orphanet J Rare Dis 2010;5:40.

4. Gatto EM, Rojas NG, Persi G, Etcheverry JL, Cesarini ME, Perandones C. Huntington disease: Advances in the understanding of its mechanisms. Clin Park Relat Disord 2020;3:100056.

5. Fodale V, Pintauro R, Daldin M, Spiezia MC, Macdonald D, Bresciani A. Quantifying Huntingtin Protein in Human Cerebrospinal Fluid Using a Novel Polyglutamine Length-Independent Assay. J Huntingtons Dis 2022;11(3):291–305.

6. Gijs M, Jorna N, Datson N, et al. High Levels of Mutant Huntingtin Protein in Tear Fluid From Huntington’s Disease Gene Expansion Carriers. J Mov Disord 2024;17(2):181–188.

7. Vauleon S, Schutz K, Massonnet B, et al. Quantifying mutant huntingtin protein in human cerebrospinal fluid to support the development of huntingtin-lowering therapies. Sci Rep 2023;13(1):5332.

8. Weiss A, Trager U, Wild EJ, et al. Mutant huntingtin fragmentation in immune cells tracks Huntington’s disease progression. J Clin Invest 2012;122(10):3731–3736.

9. Wild EJ, Boggio R, Langbehn D, et al. Quantification of mutant huntingtin protein in cerebrospinal fluid from Huntington’s disease patients. J Clin Invest 2015;125(5):1979–1986.

10. Warner JH, Long JD, Mills JA, et al. Standardizing the CAP Score in Huntington’s Disease by Predicting Age-at-Onset. J Huntingtons Dis 2022;11(2):153–171.

11. Chelsky D, Joyce C, Bockholt HJ, et al. Discovery and Targeted Proteomic Studies Reveal Striatal Markers Validated for Huntington’s Disease. Ann Clin Transl Neurol 2026;13(5):911–923.

12. Parkin GM, Corey-Bloom J, Long JD, Snell C, Smith H, Thomas EA. Associations between prognostic index scores and plasma neurofilament light in Huntington’s disease. Parkinsonism Relat Disord 2022;97:25–28.

13. Parkin GM, Corey-Bloom J, Snell C, Castleton J, Thomas EA. Plasma neurofilament light in Huntington’s disease: A marker for disease onset, but not symptom progression. Parkinsonism Relat Disord 2021;87:32–38.

14. Parkin GM, Thomas EA, Corey-Bloom J. Mapping neurodegeneration across the Huntington’s disease spectrum: a five-year longitudinal analysis of plasma neurofilament light. EBioMedicine 2024;104:105173.

15. Paulsen JS, Bovin NA, Clemsen JD, et al. Systematic Review with Meta-Analysis of Biofluid Markers for Huntington’s Disease. Mov Disord 2025;40(12):2578–2595.

16. Bockholt HJ, Clemsen JD, Baker BT, Calhoun VD, Paulsen JS. A Two-Track Model of Huntington’s Disease Pathology: Striatal Atrophy Mediates Maladaptive Immune Dysregulation. Int J Mol Sci 2026;27(5).

17. Tabrizi SJ, Scahill RI, Durr A, et al. Biological and clinical changes in premanifest and early stage Huntington’s disease in the TRACK-HD study: the 12-month longitudinal analysis. Lancet Neurol 2011;10(1):31–42.

18. Espejo EF, Guerra MD, Castellano S. Association between serum myeloperoxidase enzyme activity and Parkinson’s disease status. NPJ Parkinsons Dis 2025;11(1):94.

19. Gellhaar S, Sunnemark D, Eriksson H, Olson L, Galter D. Myeloperoxidase-immunoreactive cells are significantly increased in brain areas affected by neurodegeneration in Parkinson’s and Alzheimer’s disease. Cell Tissue Res 2017;369(3):445–454.

20. Rivera Antonio AM, Padilla M, II, Torres-Ramos MA, Rosales-Hernandez MC. Myeloperoxidase as a therapeutic target for oxidative damage in Alzheimer’s disease. J Enzyme Inhib Med Chem 2025;40(1):2456282.

21. Penney JB, Jr., Vonsattel JP, MacDonald ME, Gusella JF, Myers RH. CAG repeat number governs the development rate of pathology in Huntington’s disease. Ann Neurol 1997;41(5):689–692.

22. Langbehn DR, Brinkman RR, Falush D, Paulsen JS, Hayden MR, International Huntington’s Disease Collaborative G. A new model for prediction of the age of onset and penetrance for Huntington’s disease based on CAG length. Clin Genet 2004;65(4):267–277.

23. Zhang Y, Long JD, Mills JA, et al. Indexing disease progression at study entry with individuals at-risk for Huntington disease. Am J Med Genet B Neuropsychiatr Genet 2011;156B(7):751–763.

24. Tabrizi SJ, Schobel S, Gantman EC, et al. A biological classification of Huntington’s disease: the Integrated Staging System. Lancet Neurol 2022;21(7):632–644.

25. Huntington Study Group. Unified Huntington’s disease rating scale: Reliability and consistency. Movement Disorders 1996;11(2):136–142.

26. Mestre TA, Forjaz MJ, Mahlknecht P, et al. Rating Scales for Motor Symptoms and Signs in Huntington’s Disease: Critique and Recommendations. Mov Disord Clin Pract 2018;5(2):111–117.

27. Shoulson I, Fahn S. Huntington disease: clinical care and evaluation. Neurology 1979;29(1):1–3.

28. Long JD, Gantman EC, Mills JA, et al. Applying the Huntington’s Disease Integrated Staging System (HD-ISS) to Observational Studies. J Huntingtons Dis 2023;12(1):57–69.

29. Paulsen JS, Long JD, Johnson HJ, et al. Clinical and Biomarker Changes in Premanifest Huntington Disease Show Trial Feasibility: A Decade of the PREDICT-HD Study. Front Aging Neurosci 2014;6:78.

30. Tang C, Feigin A. Monitoring Huntington’s disease progression through preclinical and early stages. Neurodegener Dis Manag 2012;2(4):421–435.

31. Gorgolewski KJ, Auer T, Calhoun VD, et al. The brain imaging data structure, a format for organizing and describing outputs of neuroimaging experiments. Sci Data 2016;3:160044.

32. Henschel L, Conjeti S, Estrada S, Diers K, Fischl B, Reuter M. FastSurfer - A fast and accurate deep learning based neuroimaging pipeline. Neuroimage 2020;219:117012.

33. Johnson EB, Byrne LM, Gregory S, et al. Neurofilament light protein in blood predicts regional atrophy in Huntington disease. Neurology 2018;90(8):e717–e723.

34. Delporte C, Van Antwerpen P, Vanhamme L, Roumeguere T, Zouaoui Boudjeltia K. Low-density lipoprotein modified by myeloperoxidase in inflammatory pathways and clinical studies. Mediators Inflamm 2013;2013:971579.

35. Pravalika K, Sarmah D, Kaur H, et al. Myeloperoxidase and Neurological Disorder: A Crosstalk. ACS Chem Neurosci 2018;9(3):421–430.

